# COVID-19 Associated Stroke—A Single Centre Experience

**DOI:** 10.1101/2021.02.15.21249420

**Authors:** Uma Sundar, Niteen D Karnik, Amita Mukhopadhyay, Pramod Darole, Shaonak Kolte, Ashank Bansal, Yojana A Gokhale, Dnaneshwar Asole, Anagha Joshi, Sangeeta Pednekar, Swati Chavan, Trupti Trivedi, Namita Padwal, Lalana Kalekar, Charulata Londhe, Rupal Padhiyar, Dharmendra Pandey, Dhirendra Yadav, Sonal U Honrao, Prerana Bhavsar, Priyanshu Shah, Satish Gosavi, Aniket Wadal, Awesh P Shingare, Mayuri Trivedi, Gauri Pathak Oak

## Abstract

**Background and Purpose:** Various neurological complications have been reported in association with COVID-19. We report our experience of COVID-19 with stroke at a single center over a period of eight months spanning 1 March to 31 October 2020.

**Methods:** We recruited all patients admitted to Internal Medicine with an acute stroke, who also tested positive for COVID-19 on RTPCR. We included all stroke cases in our analysis for prediction of in-hospital mortality, and separately analyzed arterial infarcts for vascular territory of ischemic strokes.

**Results:** There were 62 stroke cases among 3923 COVID-19 admissions (incidence 1.6%). Data was available for 58 patients {mean age 52.6 years; age range 17–91; F/M=20/38; 24% (14/58) aged ≤40; 51% (30/58) hypertensive; 36% (21/58) diabetic; 41% (24/58) with O2 saturation <95% at admission; 32/58 (55.17 %) in-hospital mortality}. Among 58 strokes, there were 44 arterial infarcts, seven bleeds, three arterial infarcts with associated cerebral venous sinus thrombosis, two combined infarct and bleed, and two of indeterminate type. Among the total 49 infarcts, Carotid territory was the commonest affected (36/49; 73.5%), followed by vertebrobasilar (7/49; 14.3%) and both (6/49; 12.2%). Concordant arterial block was seen in 61% (19 of 31 infarcts with angiography done). ‘Early stroke’ (within 48 hours of respiratory symptoms) was seen in 82.7% (48/58) patients. Patients with poor saturation at admission were older (58 vs 49 years) and had more comorbidities and higher mortality (79% vs 38%). Mortality was similar in young strokes and older patients, although the latter required more intense respiratory support. Logistic regression analysis showed that low GCS and requirement for increasing intensity of respiratory support predicted in-hospital mortality.

**Conclusions:** We had a 1.6% incidence of COVID-19 related stroke of which the majority were carotid territory infarcts. In-hospital mortality was 55.17%, predicted by low GCS at admission.

## Introduction

Various neurological complications, ranging from hypogeusia, headache and anosmia, to Stroke, movement disorders, ataxia and myelitis, have been reported in association with COVID-19.(1,2) Acute stroke has been documented as a life threatening complication of COVID-19 in various reports since April 2020, the incidence reported ranging from 0.6% to 6%.(1–5) A pooled frequency of seven studies using the fixed effect model, estimated the incidence of acute stroke in COVID-19 to be 1.1%.(6) The proposed mechanisms include a virus induced prothrombotic state, endothelial injury, vasculitis, cardiomyopathy, microvascular thrombosis and cytokine storm.(7–9) We report our experience of COVID-19 with stroke over a period of eight months spanning March to October 2020.

Aims: We aimed to document the incidence of acute strokes among COVID-19 presentations at our hospital, to delineate the different types of strokes, and their possible mechanism. We also aimed to evaluate severity of respiratory involvement, severity of the stroke, and laboratory evidence of ongoing inflammation in these patients. Further, we aimed to determine in-hospital outcome in terms of mortality, and to evaluate the independent clinical and imaging predictors of mortality in stroke associated with COVID-19.

## Methods

Our study center is a 1400 bedded, tertiary care, municipal public hospital in Mumbai, India. We performed a prospective and retrospective observational study with serial recruitment of all patients who presented to the department of Internal Medicine with an acute stroke between 1 March and 31October 2020, who also had a reverse transcriptase-polymerase chain reaction (RT-PCR) positive report for COVID-19. Patients were diagnosed with COVID-19 using the RT-PCR tests conducted on samples collected either from a nasopharyngeal or oropharyngeal swab. We included all patients with ischemic or hemorrhagic strokes, and a combination of the two types (mixed group). We also included three patients with associated additional cerebral venous sinus thrombosis (CVST) in this study, who presented with a focal deficit and had a cerebral arterial infarct on imaging. We included patients if the presentation was with the stroke and COVID-19 was detected later during admission, as well as if the stroke developed during admission for COVID-19 related respiratory symptoms. We did not include patients with a CVST without an arterial infarct or bleed associated, in this study, as we are maintaining a separate registry of COVID-19 associated CVST at our center.

We noted the clinical, imaging and laboratory parameters in all patients, their respiratory status at admission, and the modes of supplementary oxygen that they required to maintain an oxygen saturation above 94% during hospital stay. The latter included, in order of increasing intensity, nasal cannula, venturi mask, non-rebreathing bag and mask (NRBM), high flow nasal cannula (HFNC), non-invasive ventilation (NIV), and intubation with mechanical ventilation. Two independent radiologists reviewed and opined on the images with reference to area of brain affected, vascular territory, and presence of large vessel occlusion (LVO) on CT angiography. LVO was stated to be present if Internal Carotid artery (ICA), Vertebral, Basilar, M1 or M2 segments of Middle Cerebral artery (MCA), Posterior Cerebral artery (PCA) or Anterior Cerebral artery (ACA), were occluded.

We documented the presence of existing comorbidities after checking previous medical records and ongoing medications prior to present admission. We classified patients who developed stroke symptoms either prior to or simultaneous with, or within 48 hours of respiratory symptom onset, as ‘early stroke.’

We used Microsoft Excel (2010) to tabulate and clean the raw data, and to generate pivot tables and graphs. We imported the excel datasheet into SPSS version 20.0 (Armonk, NY: IBM Corp.) and performed univariate and multivariate analyses. We reported Mean ± Standard Deviation (SD) of key continuous variables. We divided the study set into subgroups according to relevant characteristics and used the unpaired samples T-test and Fisher’s exact test to study differences in continuous and categorical variables, respectively. We reported T tests with unequal variances assumed. We calculated 95% confidence intervals for mean differences and odds ratios. We considered a p value below 0.05 to be statistically significant if the 95% confidence intervals for the concerned test statistic were congruent. We performed a binary logistic regression analysis to determine predictors of in-hospital mortality. We entered the following variables as independent predictors: sex, age, Glasgow Coma Scale (GCS) score, type of stroke (infarct, bleed, mixed infarct and bleed, infarct with associated CVST, and strokes in which the type could not be determined, classified as indeterminate), type of onset of stroke (early/late), the number of comorbidities, the number of days that the patient was symptomatic prior to admission, the patient’s oxygen saturation level on room air at the time of admission, and respiratory distress level (coded on an ascending scale reflecting the intensity of mode of supplementary oxygen delivery, where 0 represented no support required, and 6 represented invasive ventilation).

We included all the stroke cases in our analysis for prediction of in-hospital mortality, and separately analyzed arterial infarcts (pure infarcts, and the infarct component from the mixed and CVST groups) for vascular territory of ischemic strokes.

### Ethical considerations

The Human Research Institutional Ethics Committee of Lokmanya Tilak Municipal Medical College and General Hospital Mumbai, Maharashtra, India, reviewed the study and granted it scientific and ethical approval.

### Data availability statement

The data used in our analysis will be made available to researchers upon reasonable request to the corresponding author.

## Results

During the period 1 March to 31 October 2020, there were 3923 COVID-19 admissions to the medicine department at our center. Since ours is a tertiary care hospital, only patients requiring oxygen support, or with any organ dysfunction, were admitted.

We had 62 stroke admissions during this period, giving an incidence of 1.6%. We were able to collect detailed data for 58 out of these 62 patients (two patients had moved away to their village and were untraceable after discharge, and two did not revert to our communication). Table 1 shows the summarized clinical and laboratory findings for the cases. The mean age was 52.6 years (range 17–91), with 20 out of 58 patients being women. The age and sex distribution of the cases is shown in Figure 1. We had 14 patients (21% of 58) aged 40 years or less, classified as ‘young strokes.’ Comorbidities taken into account were pre-existing hypertension, diabetes mellitus, ischemic heart disease and chronic renal disease. Hypertension was present in 51% (30/58) and Diabetes in 36% (21/58) patients. Among the 58 patients, 35% (20/58) had no comorbidities, 36% (21/58) had a single comorbidity and 29% (17/58) had two or more comorbidities.

**Table 1.** COVID-19 with Stroke: Overall Data

| Parameter | Mean±SD | Range |
| --- | --- | --- |
| Age | 52.6±15.47 | 17–91 |
| GCS score | 9.31±4.08 | 3–15 |
| SpO2 at admission | 89.16±13.30 | 46–99 |
| Platelet count/μL (n=53) | 220718±106107 | 20100–607000 |
| CRP mg/L (n=31) | 61.88±58.23 | 2.8–229 |
| D Dimer ng/mL (n=27) | 2796.3±2389.6 | 335–10000 |
| D Dimer nmol/L (n=27) | 30.62±26.17 | 3.67–109.52 |
| S. Creatinine mg/dL (n=54) | 1.22±0.70 | 0.4–5.2 |
| S. Creatinine μmol/L (n=54) | 107.88±61.47 | 35.36–459.68 |
| Gender ratio M/F (%) | 38/20 (66:34) |  |
| Pts with $\geq 1$ comorbidity (%) | 38/58 (65) | |
| Infarcts/Pure Bleeds (% of 58) | 49/7 (84/12) |  |
| Carotid/Vertebrobasilar/Both (%) | 36/7/6 (73.4/14.3/12.3) |  |
| Proximal arterial block (%) | 19/31 (61) |  |
| SpO<sub>2</sub> < 95% at admission (%) | 24/58 (41) |  |
| Mortality (%) | 32/58 (55.17) |  |

CRP: C Reactive Protein

**Figure 1.**
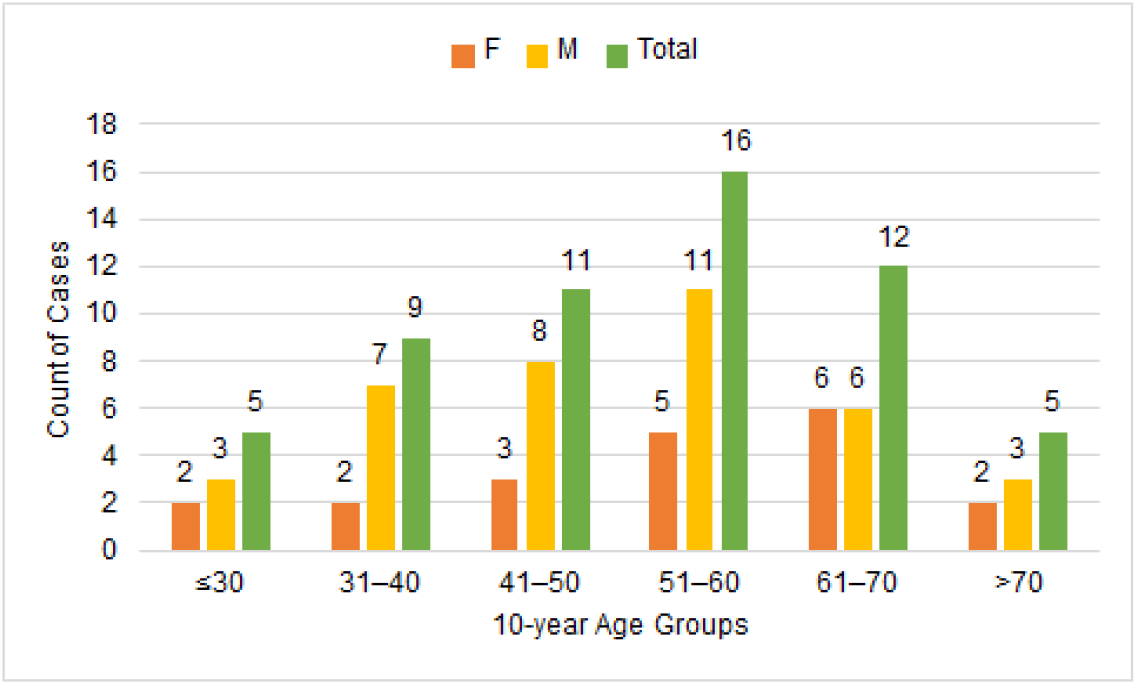
Age-Sex Distribution of COVID Stroke Cases

Among 58 strokes, 44 were pure arterial infarcts, seven were arterial bleeds, two were mixed (arterial infarct plus bleed), three had associated cerebral venous sinus thrombosis (CVST) with cortical infarcts in the carotid territory (a total of 49 out of 58 patients thus had arterial infarcts), and two were indeterminate (Figure 2A). These last were both bithalamic infarcts, possibly due to blockage of artery of Percheron or vein of Galen. As CT angiography and venography were not available for these two patients, they have been classified as of indeterminate etiology.

**Figure 2.**
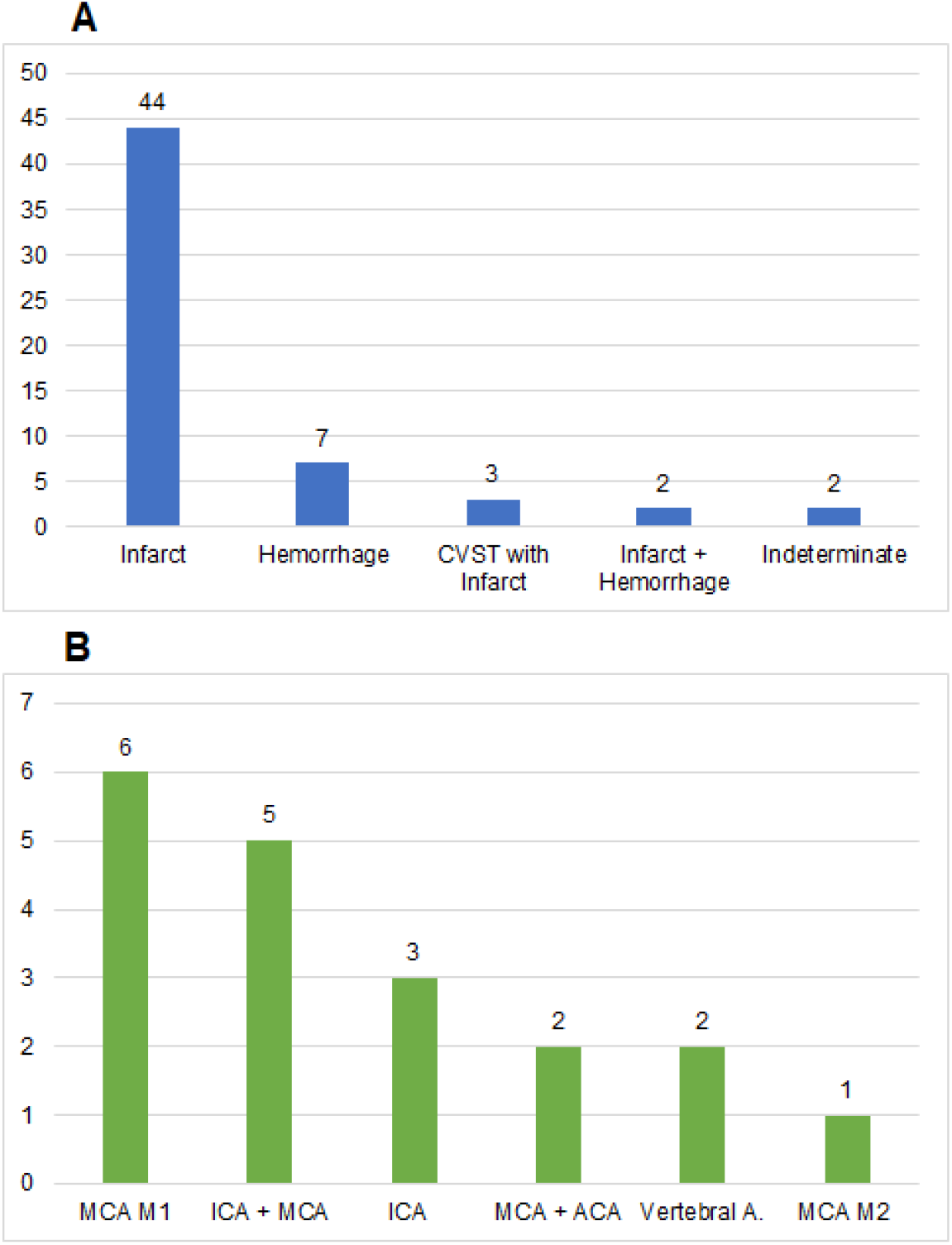
A: Types of Stroke in COVID Stroke Patients and 2B: Large Vessel Occlusion Types among Arterial Infarcts. CVST: Cerebral Venous Sinus Thrombosis; MCA M1: M1 segment of Middle Cerebral artery; ICA: Internal Carotid artery; ACA: Anterior Cerebral artery; MCA M2: M2 segment of Middle Cerebral artery. CT Angiographies done in 31 of whom 19 had concordant arterial blocks.

Among the 49 patients with ischemic strokes either in isolation or in combination with CVST or arterial bleed, 36 patients (36/49; 73.5%) had infarcts in the carotid territory, seven (7/49; 14.3%) in the vertebrobasilar territory, and six patients (6/49; 12.2%) had infarcts in both territories. Among the carotid ischemic strokes (total 42), 39 scans were available for analysis for type of carotid territory infarct. Among these, 16 were large hemispherical, 10 were subcortical and 13 were pure cortical. A concordant arterial block was seen in 61% (19/31) of the infarcts which had angiography done. The commonest vessels occluded were MCA main stem (6/19), ICA extending into MCA (5/19), and ICA alone (3/19). (Figure 2B)

In this series, 82.7% (48/58) patients had an ‘early stroke,’ defined as development of stroke symptoms either prior to or simultaneous with, or within 48 hours of respiratory symptom onset. We noted 22/58 patients (37.9%) who presented to the emergency room with the stroke, with no other associated respiratory or systemic symptoms. A comparative analysis of patients with early versus late stroke, is shown in Supplementary Table S1.

Sixteen of 58 (28%) patients had already had over 72 hours of respiratory symptoms at home before presentation. Six of these patients had stroke symptoms in addition, at home, but still presented 72 hours or later after their first symptom. On analysis of these six patients, four had a lacunar syndrome and four had GCS between 9 and 13 at admission.

In this series, 34/58 (58.6%) patients had normal oxygen saturation on room air (95% and above) at presentation. There were 22/58 patients (37.9 %) with no respiratory symptoms either, at presentation, with normal saturation on room air in casualty. Among the 34 patients who had good oxygen saturation at admission, 22 continued to remain on room air, four went on to nasal cannula at 4–5 liters/minute, one received venturi mask support at 8 liters/minute, one through NRBM at 15 liters/minute, four received oxygen through NIV with average positive end-expiratory pressure (PEEP) at 7 cm water, and two required intubation and ventilation. Among the 24 patients who had poor oxygen saturation at admission, eight went on to nasal cannula at 4–5 liters/minute, five received venturi mask support at 8 liters/minute, one through NRBM at 15 liters/minute, one through HFNC at up to 60 liters/minute, eight received oxygen through NIV at average PEEP 7 cm water, and one required intubation and ventilation. Mortality in the former group was 38.2% and in the latter, 79.1%; a significant difference. Comparison between these two groups (Table 2) showed that patients in the group with poor saturation at admission were significantly older, and fewer of them were free of comorbidities (20.8% vs 44.1% of patients with good saturation; although this was not statistically significant).

**Table 2.**
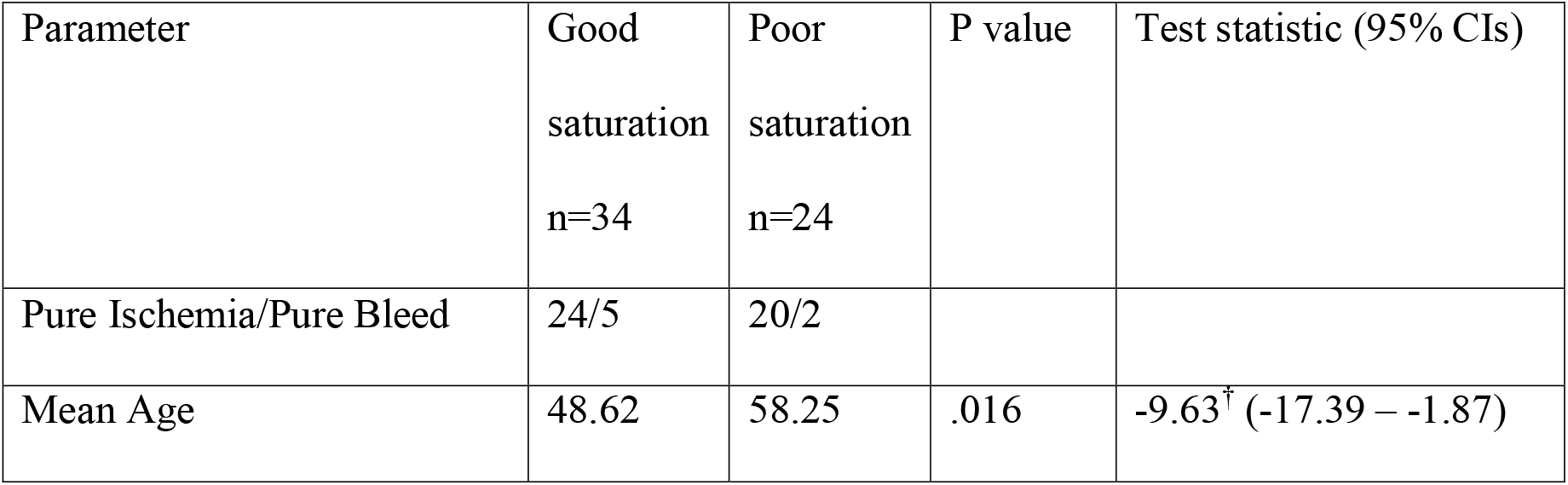
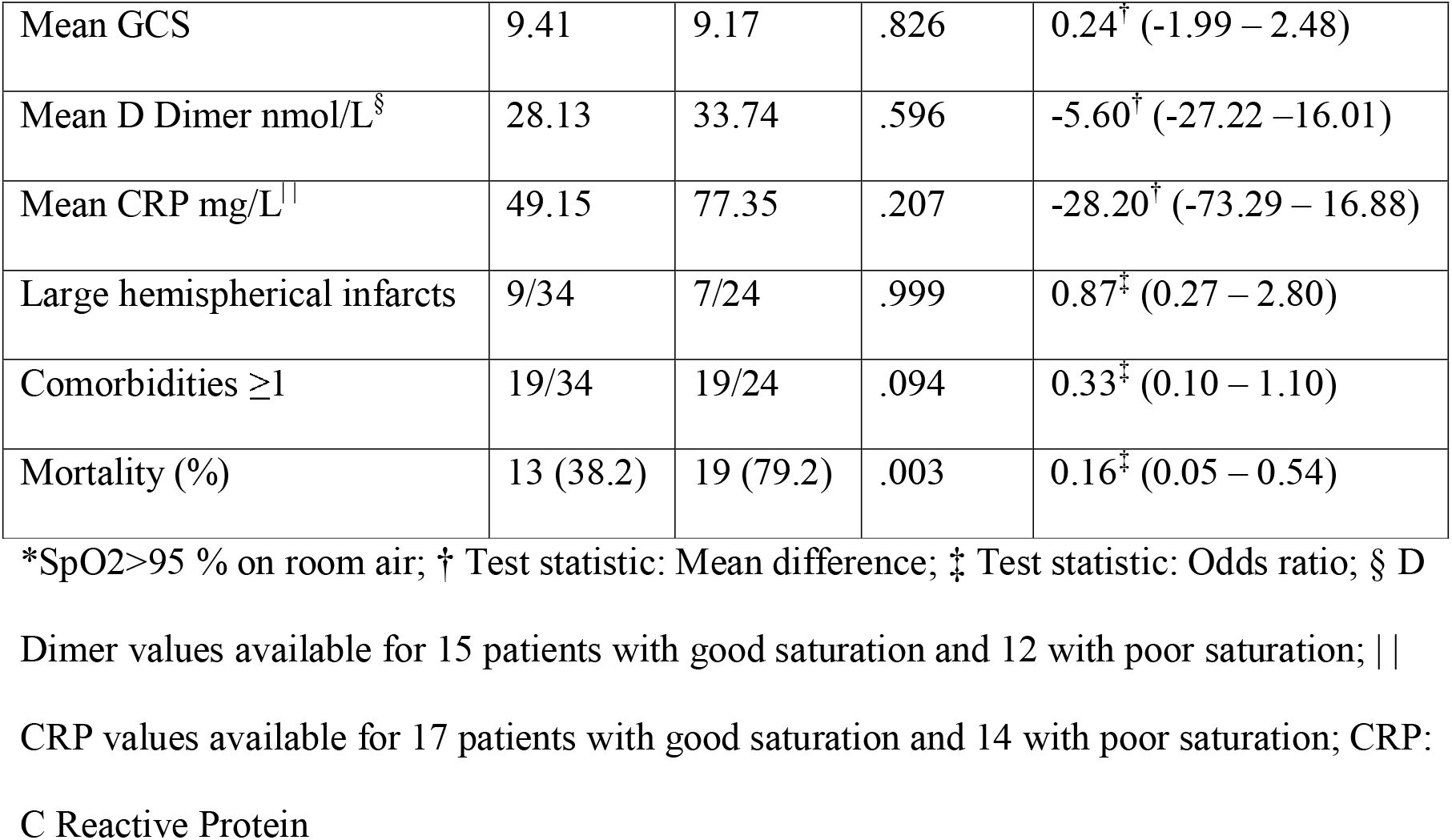
COVID-19 associated stroke patients with good saturation* at admission compared to those with low saturation.

Compared to younger patients, 40 years and under (24%; 14/58), older patients with stroke in COVID-19 did not have notably higher mortality (Table 3). More of the older patients, as expected, had associated comorbidities. Concordant arterial block (analyzed in a subset of 31 patients who had CT angiography reports available), was significantly commoner in the younger group (100% in younger group vs 43% in older group). A larger proportion of the younger patients, as compared to the older group, had normal oxygen saturation at admission. In fact, 13/14 (92.8%) ‘young strokes’ did not progress beyond low flow systems (up to venturi mask) in supplementary oxygen requirements during admission, whereas 17/44 (38.6%) older patients required respiratory support with NRBM, HFNC, NIV, or intubation with mechanical ventilation. In contrast, the mortality was similar in both the groups (50% in young strokes vs 56.8% in older strokes).

**Table 3.**
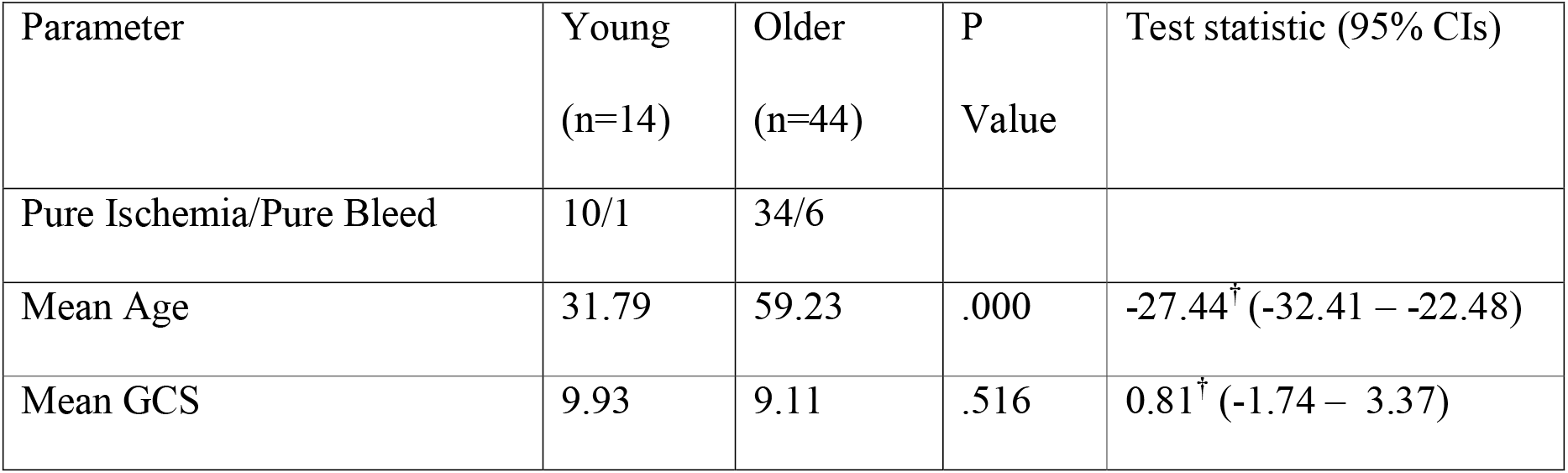
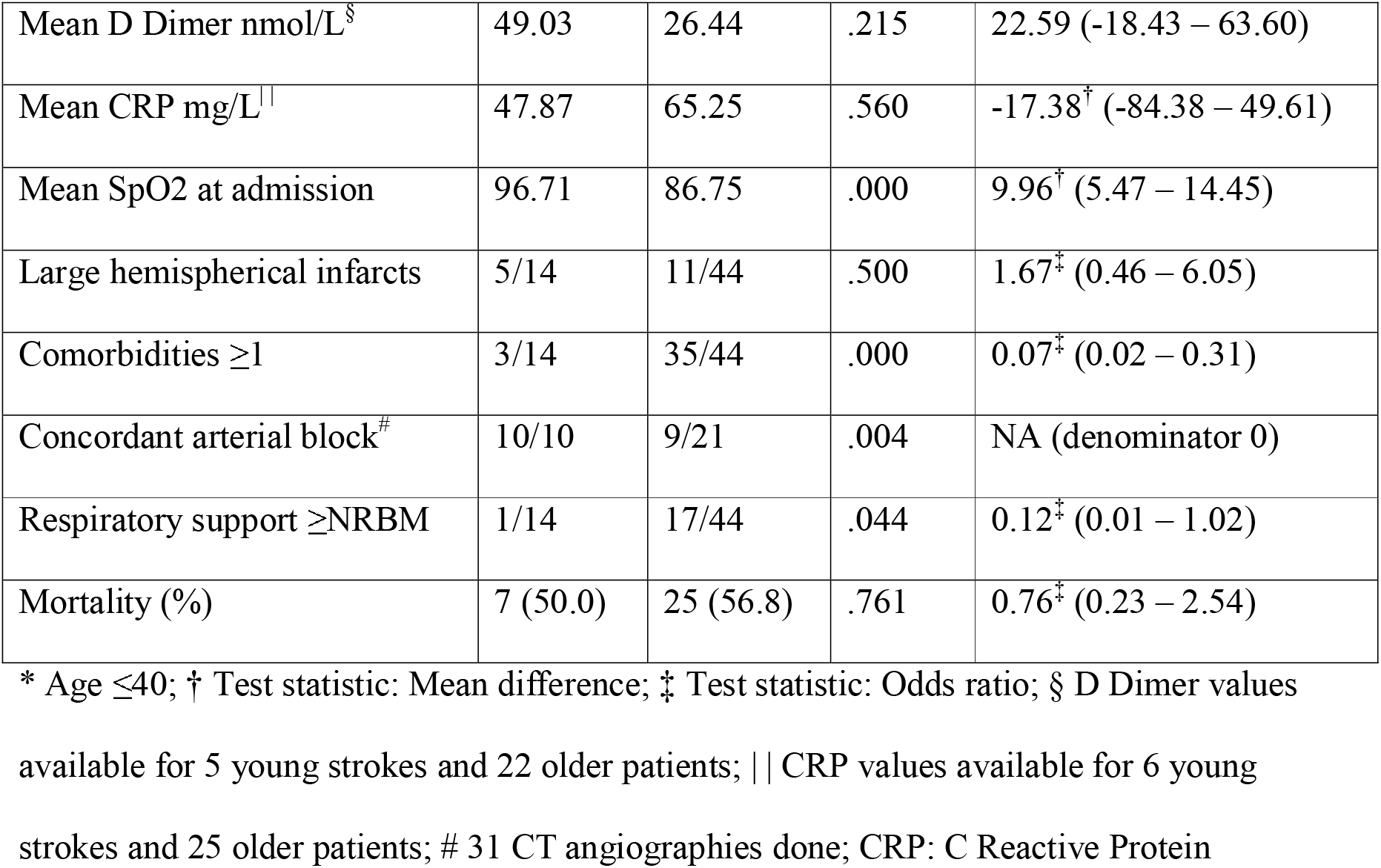
Young strokes* in COVID-19: a comparison with older strokes

| Parameter | Young<br>(n=14) | Older<br>(n=44) | P<br>Value | Test statistic (95% CIs) |
| --- | --- | --- | --- | --- |
| Pure Ischemia/Pure Bleed | 10/1 | 34/6 |  |  |
| Mean Age | 31.79 | 59.23 | .000 | -27.44<sup>†</sup> (-32.41 – -22.48) |
| Mean GCS | 9.93 | 9.11 | .516 | 0.81<sup>†</sup> (-1.74 – 3.37) |
| Mean D Dimer nmol/L<sup>§</sup> | 49.03 | 26.44 | .215 | 22.59 (-18.43 – 63.60) |
| Mean CRP mg/L | 47.87 | 65.25 | .560 | -17.38<sup>†</sup> (-84.38 – 49.61) |
| Mean SpO2 at admission | 96.71 | 86.75 | .000 | 9.96<sup>†</sup> (5.47 – 14.45) |
| Large hemispherical infarcts | 5/14 | 11/44 | .500 | 1.67<sup>‡</sup> (0.46 – 6.05) |
| Comorbidities ≥1 | 3/14 | 35/44 | .000 | 0.07<sup>‡</sup> (0.02 – 0.31) |
| Concordant arterial block<sup>#</sup> | 10/10 | 9/21 | .004 | NA (denominator 0) |
| Respiratory support ≥NRBM | 1/14 | 17/44 | .044 | 0.12<sup>‡</sup> (0.01 – 1.02) |
| Mortality (%) | 7 (50.0) | 25 (56.8) | .761 | 0.76<sup>‡</sup> (0.23 – 2.54) |

\* Age ≤ 40; † Test statistic: Mean difference; ‡ Test statistic: Odds ratio; § D Dimer values
available for 5 young strokes and 22 older patients; || CRP values available for 6 young
strokes and 25 older patients; # 31 CT angiographies done; CRP: C Reactive Protein

In our series, 47/58 patients were admitted after the beginning of July, when Remdesivir injections became available to us. Patients with significant hypoxia, within 10 days of onset of the first symptom of COVID-19, and with a normal creatinine clearance, qualified for Remdesivir therapy. In this series, 19 of 47 patients received Remdesivir. Similarly, Tocilizumab was available to us from early May (52 patients were admitted after this point), and was administered to patients with a diagnosis of cytokine storm, as judged by persistent hypoxia with diffuse lung involvement on CT chest, and raised inflammatory markers. In this series, 7 of 52 patients received 400 mg of intravenous Tocilizumab. However, neither Remdesivir nor Tocilizumab showed correlation with in-hospital survival in this series (Supplementary Tables S2 and S3 respectively).

Figure 3 shows the timeline of stroke admissions in our series, in relation to overall COVID-19 admissions from March to October 2020. Whereas there was a fall in COVID-19 cases and flattening of the curve from August onwards, the maximum number of stroke admissions (32/58) were in September and October 2020.

**Figure 3:**
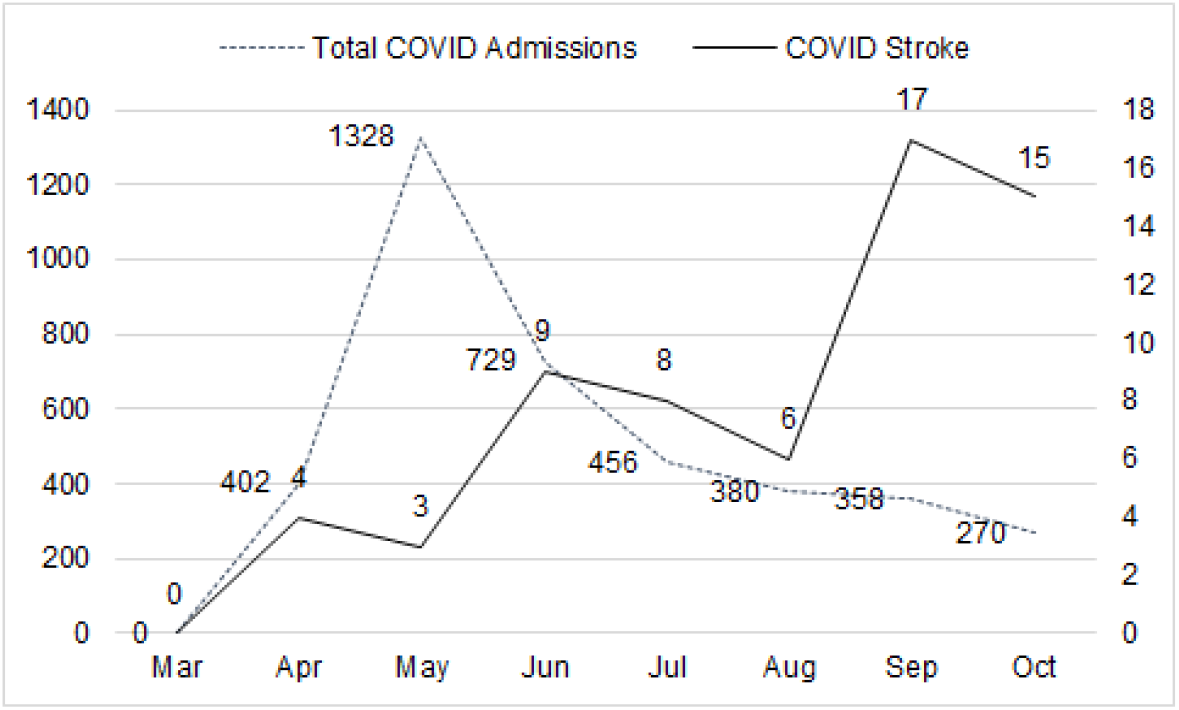
Time Trend of COVID Stroke Cases vs Total COVID-19 Admissions to Internal Medicine

In-hospital mortality rate was 32/58 (55.17 %). Mortality among younger patients (≤ 40 yrs) was 7/14 (50%). Binary logistic regression (Table 4) showed level of need for respiratory support and GCS at admission to be independent predictors of mortality. Progression to more intense modes of support correlated positively with mortality, and higher GCS at admission correlated negatively. “Early Stroke’ did not achieve significance in relation to mortality in binary or multivariate analyses.

**Table 4.**
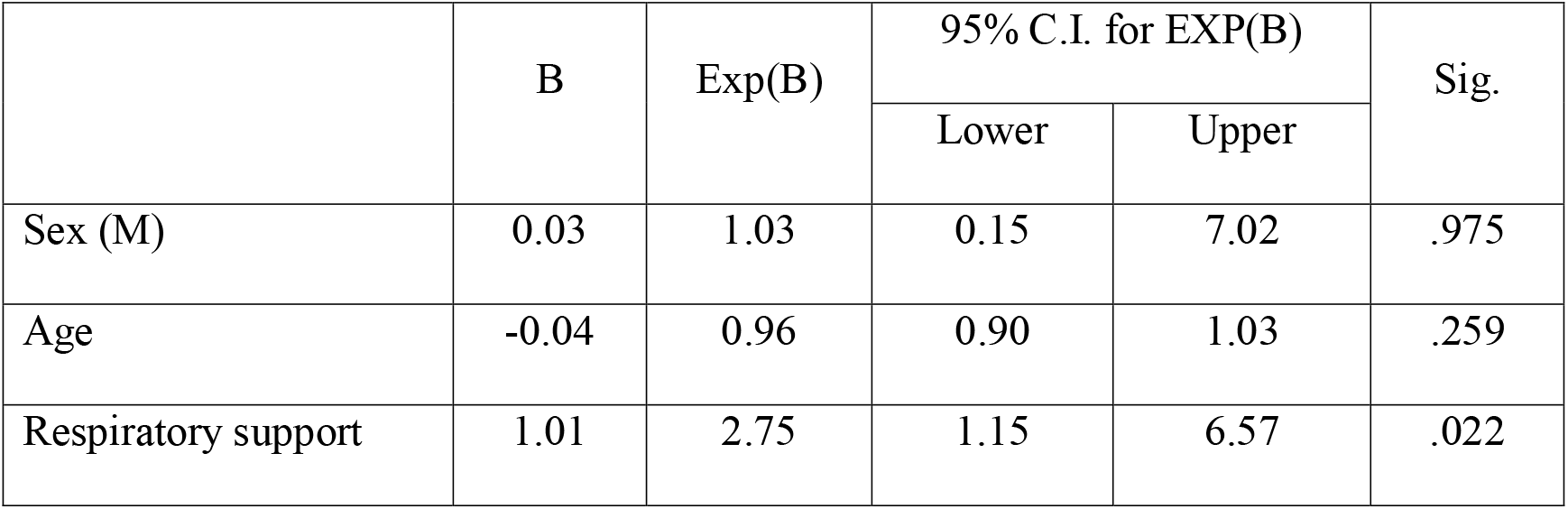
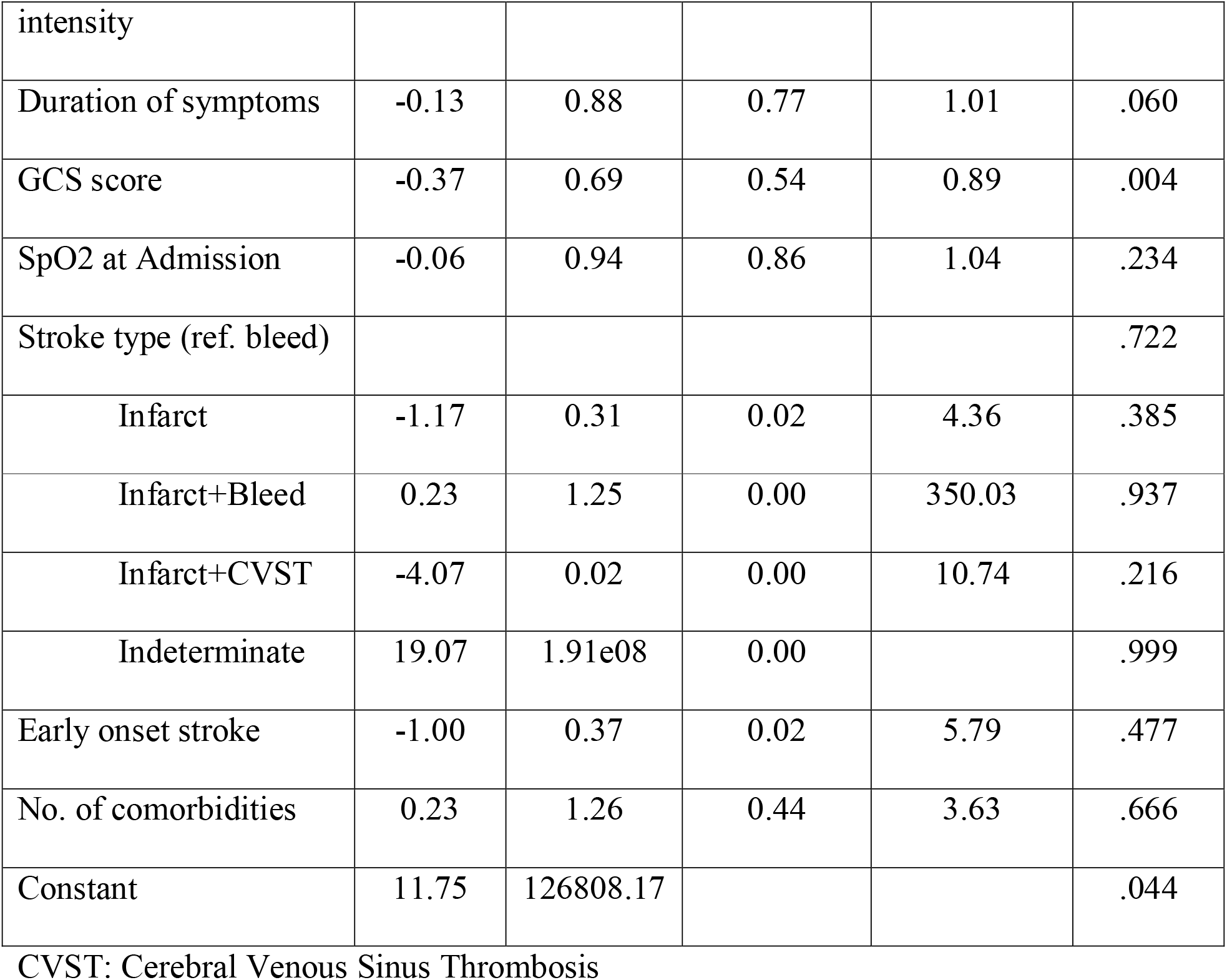
Predictors of In-hospital Mortality in COVID-19

## Discussion

Our incidence of acute stroke during COVID-19 is 1.6%. However, this is probably an underestimation. In the months of March and April 2020, we did not recognize that stroke could be the first presentation of COVID-19. Hence, if the systemic or respiratory involvement was mild or absent in COVID related strokes in the early months, the cases could have been missed. Additionally, logistical problems of testing in the early months contributed to the underestimation. We have not included cerebral venous sinus thrombosis in this study, as mentioned, except in three cases where CVST was found on imaging in association with a carotid territory infarct. The incidence of stroke in COVID-19 in other reported studies ranged from 0.6% to 6%.(1–5,10–17) This wide range of reported incidence is due to differing nature of data collection at different centers, with some studies reporting only specific subtypes of strokes, and some others considering only severe cases of COVID-19 for evaluation.

We classified the strokes as presenting early, or late, in relation to the first systemic symptom of fever or respiratory complaints. This was to estimate the proportion of patients who had a neurological presentation of COVID-19. In our series, 83% of patients with stroke had the neurological symptoms within 48 hours of the first systemic/respiratory complaint. In fact, 22 patients (22/58; 37.9 %) had a pure neurological presentation, with no respiratory or systemic symptoms at admission. This very early thromboembolic or hemorrhagic presentation argues for a high thrombo-inflammatory pathology in these patients. In a meta-analysis of 28 articles related to stroke and COVID-19, the average number of days to develop stroke among patients after the onset of COVID-19 infection was 6.9 ± 4.5 days.(6) Some series, similar to our data, have reported that patients may present with stroke with simultaneous asymptomatic COVID-19 infection.(11,17) In concordance with stroke data in non-COVID-19 times, and with data published on stroke in COVID-19, the vast majority of our CVAs were infarcts.(13,15–17) The majority were in the Carotid territory. Over two thirds of the patients who had an angiography done, showed concordant arterial blocks in the large vessels, MCA main stem being the commonest vessel occluded. In the meta-analysis by Wei Lee et al.(6) and in various studies,(3,11,17) occluded vessels involved were M1 vessels, M2 vessels, internal carotid, multi-territorial, posterior cerebral, basilar, ACA, and the vertebral artery. Mohamud et al have also raised the issue of Internal Carotid thrombosis in COVID-19 associated stroke being linked to cytokine storm.(18)

Although echocardiography is a mandatory investigation in acute ischemic stroke, we could not perform this test in our patients due to logistical problems of transporting COVID-19 patients for tests, and not having a dedicated machine for echocardiography in the COVID-19 wards. We plan to do echocardiography in stable patients on follow up, after they are COVID-19 negative. The elucidation of mechanism of ischemic stroke requires determination of area of infarct, followed by, if required, angiography, echocardiography, prolonged Holter monitoring and tests of coagulation if deemed necessary. During the COVID-19 pandemic, it is not possible to do many of these tests in a resource challenged setting, and the stroke may hence be labelled ‘cryptogenic,’ as investigations are limited. Yaghi et al, comparing COVID positive and COVID negative strokes, reported a higher incidence of both LVO and cryptogenic strokes among the former.(13)

Among our patients, 24% were 40 years of age or under. They had significantly fewer comorbidities and higher oxygen saturation at admission, as compared to the older group. Mortality was, however, similar in both groups, possibly contributed to by the significantly higher proportion of proximal vessel occlusion in the younger group (100% in the young group, vs 43% in the older group, in the patients who had a CT angiography done). This could have led to rapidly worsening stroke and subsequent mortality, despite mild respiratory illness. This is borne out by the fact that 13/14 of these young strokes had a mild respiratory involvement, never requiring supplementary oxygen beyond venturi mask support. It is possible that some of these young patients had tobacco or alcohol abuse as a risk factor for stroke in the background. We do not have uniform data on tobacco and alcohol addiction among our patients, hence these were not considered in the analysis. Various studies have noted the association of COVID-19 with stroke in the young.(6,11,17,19) Belani et al have reported a 7-fold increase in the rate of large vessel stroke in young people compared with the previous year, in New York city, associated with a hypercoagulable state, due to COVID-19.(20)

On comparing the patients who were admitted with a normal oxygen saturation with those who had a low saturation at admission, we found significantly more patients with higher mean age in the latter group, possibly leading to the higher mortality in this group. However, subtype of stroke, level of inflammatory markers and presence of comorbidities were similar in both groups. (Table 2)

Although a large proportion of our patients presented after both Remdesivir and Tocilizumab became available to us for treatment, the addition of these did not appear to improve survival. Remdesevir, which has been shown to reduce hospitalization time in COVID-19 with lower respiratory involvement,(21) is unlikely to have much effect on the course of stroke in COVID-19, which is multifactorial and complex in etiopathogenesis. Tocilizumab, which binds specifically to both soluble and membrane-bound interleukin-6 receptors, is used in the management of COVID-19 associated cytokine storm; however, whether it independently results in survival benefit in severe COVID-19, is a matter for debate.(22,23) Although cytokine storm has been proposed as an etiological factor in stroke in COVID-19, it is only one of many proposed factors.

Since our hospital admits only the patients requiring a higher level of care, the curve of overall COVID-19 admissions specifically reflects the most severe cases only (Figure 3). Within this category, it is clear that the incidence of stroke increased as the number of severe admissions fell, during August to October 2020. The explanation for this is unclear—it is possible that the virus had a higher thromboinflammatory effect during this period, or possibly some of the patients may have been reinfected after a previous mild illness and recovery; patients and relatives could also have been more amenable to coming for admission for stroke as the number of cases declined overall in the city.

In this series, 55% of patients expired in hospital. We found lower GCS score at admission to be an independent predictor of in-hospital mortality in stroke with COVID-19, as expected, signifying a worse stroke. Requirement for higher levels of respiratory support also predicted mortality, signifying more severe respiratory and neurological illness overall. Early onset of stroke, longer duration of symptoms at home before presenting to hospital, and oxygen saturation at admission showed no correlation with in-hospital mortality. In three studies comparing stroke in COVID-19 patients with stroke in non-COVID-19 patients, mortality was higher in all three cohorts among patients with COVID-19 infection; Benussi et al.(16) reporting 34.9% vs. 5.9%, Yaghi et al.(13) reporting 63.6% vs. 9.3%, and Escalard et al.(24) reporting 60.0% vs. 11.0%, respectively. Taylor et al reported a mortality of 46% among their patients, with a high percentage of proximal LVOs.(25)

We faced many management dilemmas during the treatment of these 58 patients. In the initial months, patients would not be brought on time, or not brought at all to hospital, due to fear of COVID-19 among relatives. Although six patients were brought in the golden period for intravenous thrombolysis, and one developed ischemic stroke while in the ward, intravenous thrombolysis was not possible in any of them due to various factors—severe respiratory disease, or severe stroke, and in one case, absence of relatives to sign consent, due to quarantine. We detected only a single patient with platelet count below 50000/ c mm during admission, in our study. Hence, apprehensions about thrombocytopenia should probably not hinder the decision to thrombolyse immediately, if the patient meets criteria for intravenous thrombolysis.

We follow a policy of anticoagulation with Heparin during admission, in all patients with moderate to severe COVID-19 requiring oxygen supplementation, who have raised d Dimer over twice normal levels. Similarly, we discharge moderate to severe COVID-19 patients on oral anticoagulants, to be taken for 4-6 weeks, to prevent thromboembolic episodes, in line with current guidelines.(26,27) In the presence of a large infarct with attendant advanced age or uncontrolled hypertension, there is a risk of a secondary bleed into the infarct while on Heparin. We decided the dose and type of Heparin during hospital stay, and the oral anticoagulant at discharge, on a case to case basis. We prescribed a single antiplatelet at discharge in addition to the oral anticoagulant, in the majority of our patients. Treatment of COVID positive ischemic stroke, in the available literature, has ranged from mechanical thrombectomy in very stable patients, to standard oral antiplatelet therapy,(3,24,28,29) reflecting the wide variability in severity of stroke as well as respiratory illness, in them. In conclusion, we had an incidence of 1.6%, of COVID-19 related stroke in our center. Of these 84% were infarcts, with the majority being in the Carotid territory. Young strokes comprised 24% of the patients. Concordant arterial occlusion was seen in 61% of the patients who had an angiography done, and was significantly more frequent in younger patients compared to older patients, in our study group. In-hospital mortality was 55.17% and was predicted by a low GCS at admission and higher levels of respiratory support required to maintain saturation. Management dilemmas included the role of anticoagulation during admission and at discharge, taking into account the importance of preventing further thromboembolic complications vs the risk of secondary bleeding into large infarcts.

## Supporting information

STROBE checklist

IEC approval document

## SUPPLEMENTARY MATERIAL

### Supplementary Tables

**Table S1.**
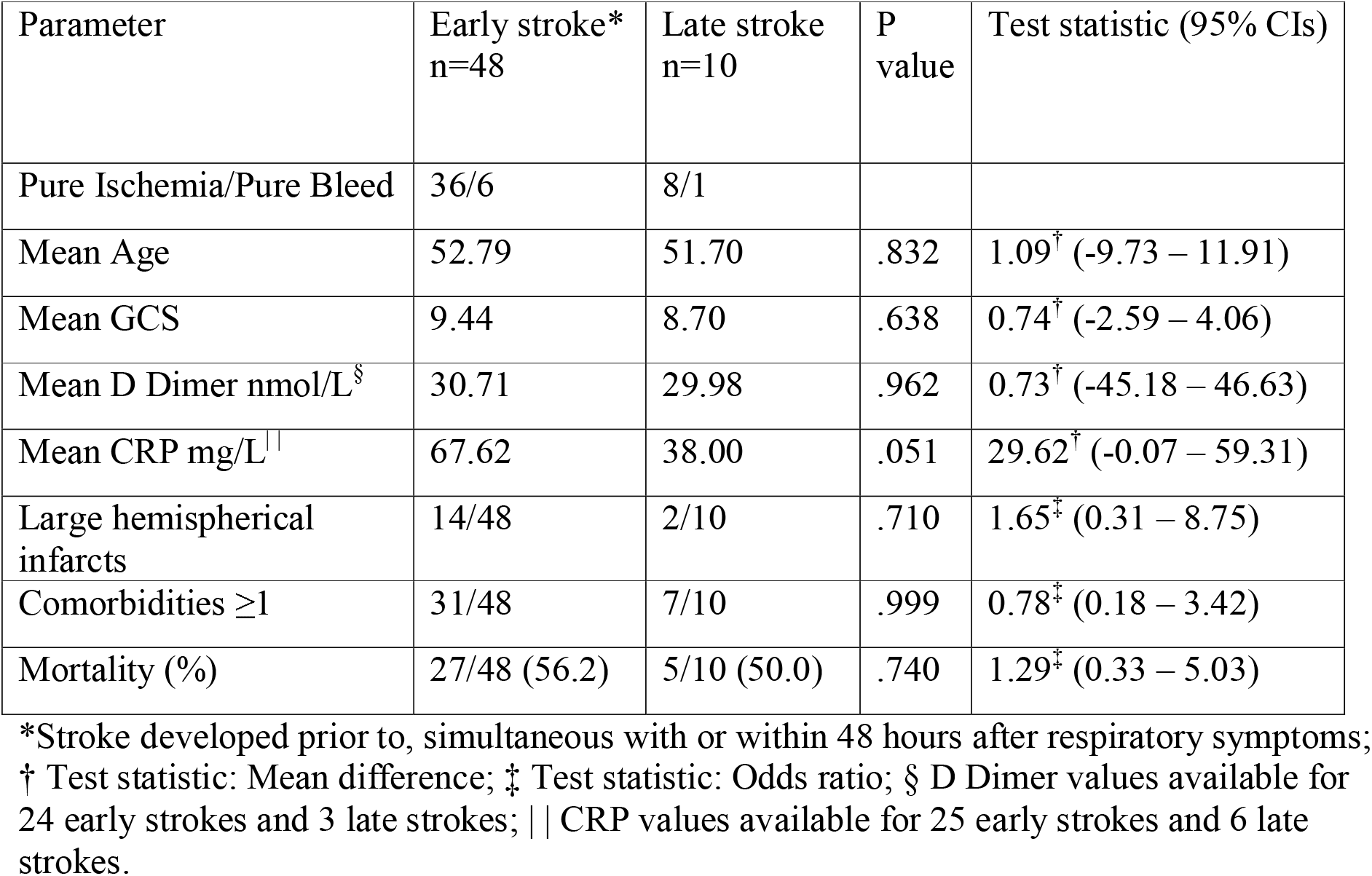
COVID-19 associated stroke patients with early stroke vs. those with late stroke

| Parameter | Early stroke*<br>n=48 | Late stroke<br>n=10 | P<br>value | Test statistic (95% CIs) |
| --- | --- | --- | --- | --- |
| Pure Ischemia/Pure Bleed | 36/6 | 8/1 |  |  |
| Mean Age | 52.79 | 51.70 | .832 | 1.09<sup>†</sup> (-9.73 – 11.91) |
| Mean GCS | 9.44 | 8.70 | .638 | 0.74<sup>†</sup> (-2.59 – 4.06) |
| Mean D Dimer nmol/L<sup>§</sup> | 30.71 | 29.98 | .962 | 0.73<sup>†</sup> (-45.18 – 46.63) |
| Mean CRP mg/L | 67.62 | 38.00 | .051 | 29.62<sup>†</sup> (-0.07 – 59.31) |
| Large hemispherical<br>infarcts | 14/48 | 2/10 | .710 | 1.65<sup>‡</sup> (0.31 – 8.75) |
| Comorbidities $\geq 1$ | 31/48 | 7/10 | .999 | 0.78<sup>‡</sup> (0.18 – 3.42) |
| Mortality (%) | 27/48 (56.2) | 5/10 (50.0) | .740 | 1.29<sup>‡</sup> (0.33 – 5.03) |

\*Stroke developed prior to, simultaneous with or within 48 hours after respiratory symptoms;
<sup>†</sup> Test statistic: Mean difference; <sup>‡</sup> Test statistic: Odds ratio; <sup>§</sup> D Dimer values available for 24 early strokes and 3 late strokes; <sup>||</sup> CRP values available for 25 early strokes and 6 late strokes.

**Table S2.** Analysis of patients admitted after Remdesivir became available

| Parameter | Received Rem*, n=19 | Did not receive Rem, n=28 | P value | Test statistic (95% CIs) |
| --- | --- | --- | --- | --- |
| Pure Ischemia/Pure Bleed | 15/2 | 19/4 |  |  |
| Mean Age | 54.95 | 51.79 | .510 | 3.16<sup>†</sup> (-6.45 – 12.77) |
| Mean SpO2 at admission | 87.16 | 90.54 | .427 | -3.38<sup>†</sup> (-11.88 – 5.13) |
| Mean GCS | 9.42 | 9.32 | .935 | 0.10<sup>†</sup> (-2.36 – 2.56) |
| Mean D Dimer nmol/L§ | 29.57 | 32.16 | .806 | -2.59<sup>†</sup> (-24.22 – 19.04) |
| Mean CRP mg/L\|\| | 58.18 | 66.38 | .705 | -8.21<sup>†</sup> (-52.23 – 35.81) |
| Mean Creatinine µmol/L# | 102.36 | 120.36 | .317 | -18.00<sup>†</sup> (-54.01 – 18.00) |
| Large hemispherical infarcts | 4/19 | 8/28 | .737 | 0.67<sup>‡</sup> (0.17 – 2.63) |
| Comorbidities ≥1 | 10/19 | 19/28 | .365 | 0.53<sup>‡</sup> (0.16 – 1.75) |
| Mortality (%) | 10/19<br>(52.6%) | 14/28<br>(50.0%) | .999 | 1.11<sup>‡</sup> (0.35 – 3.57) |

\*Remdesivir available for use post July 1, n = 47; <sup>†</sup> Test statistic: Mean difference; <sup>‡</sup> Test statistic: Odds ratio; § D Dimer values available for 16 patients who received Remdesivir and 11 who did not; || CRP values available for 17 patients who received Remdesivir and 14 who did not; # Creatinine values available for 19 patients who received Remdesivir and 26 who did not.

**Table S3.** Analysis of patients admitted after Tocilizumab became available

|  | Received<br>Toci*, n=7 | Did not<br>receive<br>Toci, n=45 | P<br>value | Test statistic (95% CIs) |
| --- | --- | --- | --- | --- |
| Pure Ischemia/Pure Bleed | 6/0 | 32/7 |  |  |
| Mean Age | 67.00 | 50.13 | .005 | 16.87<sup>†</sup> (6.40 – 27.33) |
| Mean SpO2 at admission | 82.43 | 90.76 | .117 | -8.33<sup>†</sup> (-19.21 – 2.56) |
| Mean GCS | 9.29 | 9.09 | .909 | 0.20<sup>†</sup> (-3.62 – 4.02) |
| Mean D Dimer nmol/L§ | 27.15 | 31.84 | .605 | -4.69<sup>†</sup> (-23.28 – 13.91) |
| Mean CRP mg/L\|\| | 93.17 | 54.38 | .260 | 38.79<sup>†</sup> (-36.99 – 114.58) |
| Mean Creatinine µmol/L# | 1.36 | 1.22 | .520 | 12.04<sup>†</sup> (-27.41 – 51.49) |
| Large hemispherical<br>infarcts | 2/7 | 12/45 | .999 | 1.10<sup>‡</sup> (0.19 – 6.44) |
| Comorbidities ≥1 | 6/7 | 26/45 | .228 | 4.38<sup>‡</sup> (0.49 – 39.50) |
| Resp. support ≥NRBM | 5/7 | 13/45 | .041 | 6.15<sup>‡</sup> (1.06 – 35.84) |
| Mortality (%) | 3/7 (42.9%) | 24/45<br>(53.3%) | .698 | 0.66<sup>‡</sup> (0.13 – 3.27) |

\*Tocilizumab available for use post May 13, n = 52; † Test statistic: Mean difference; ‡ Test statistic: Odds ratio; § D Dimer values available for 7 patients who received Tocilizumab and 20 who did not; || CRP values available for 6 patients who received Tocilizumab and 25 who did not; # Creatinine values available for 7 patients who received Tocilizumab and 43 who did not.

