## Supplementary figures and images for "COVID-19 Associated Stroke—A Single Centre Experience"

### IEC approval document

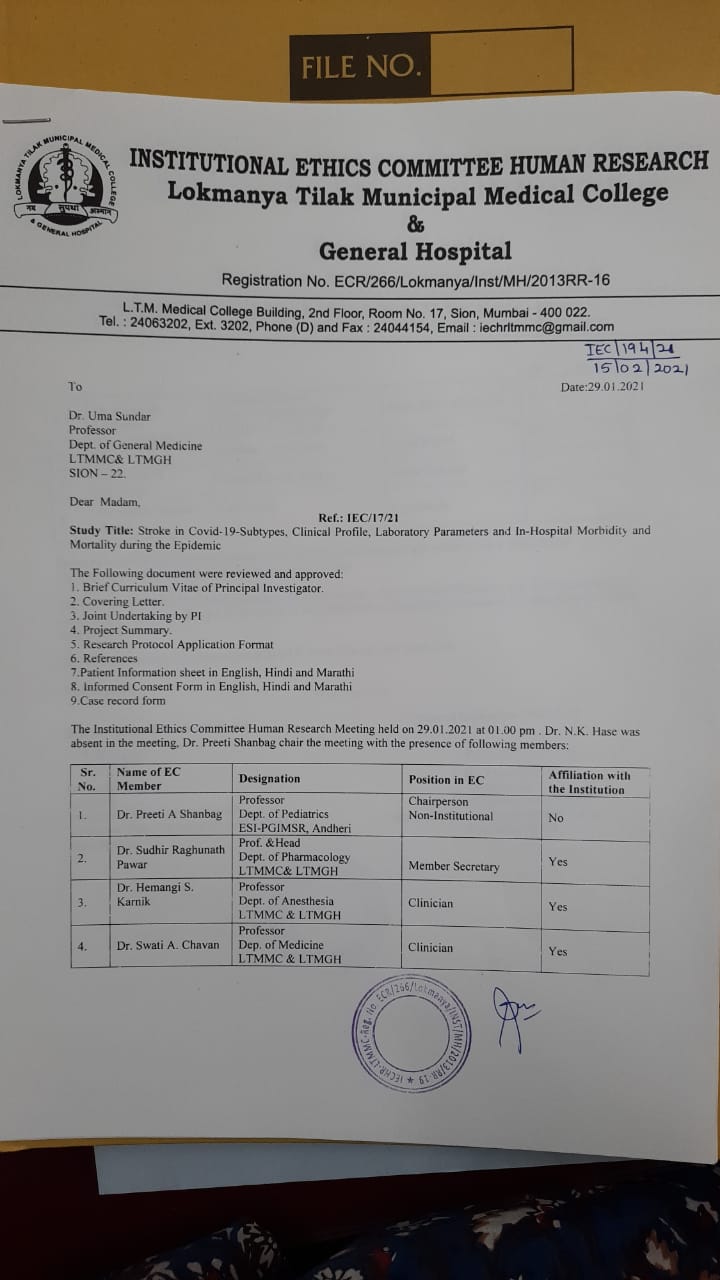

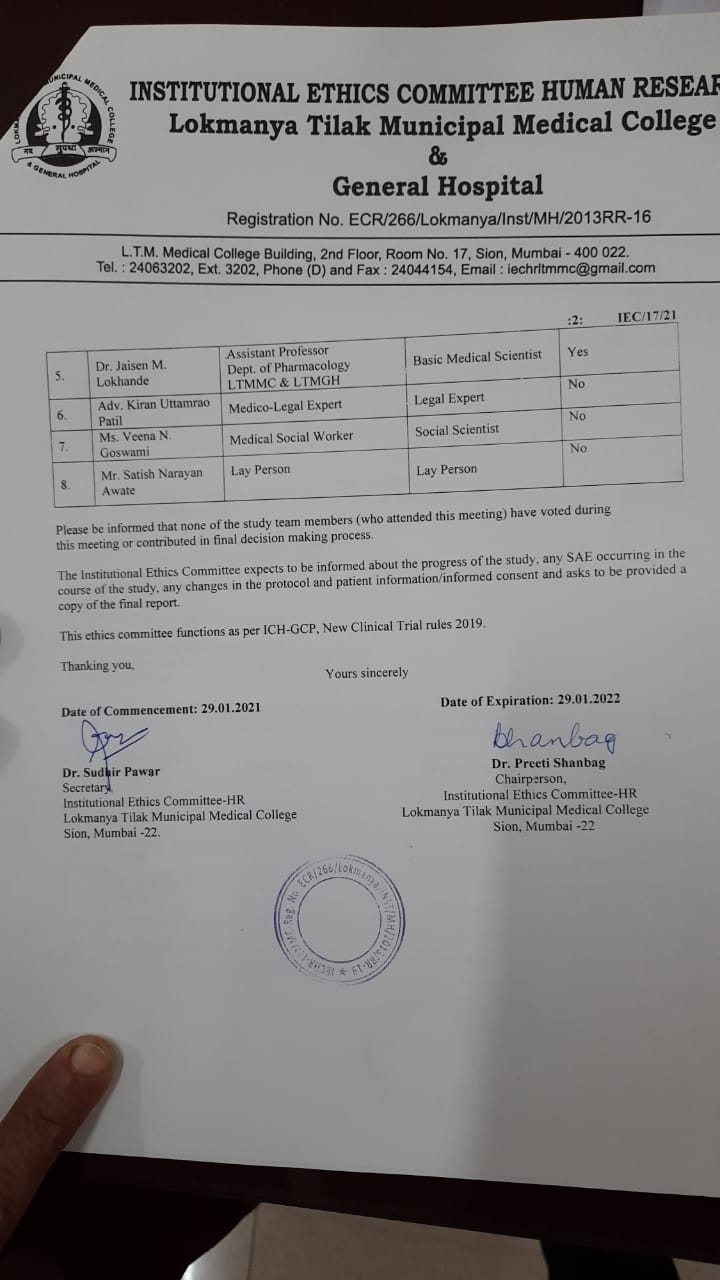
